# Antidepressants and Slower Disease Progression in Huntington’s Disease

**DOI:** 10.1101/2025.03.18.25323225

**Authors:** Duncan Mclauchlan, Cheney Drew, Peter Holmans, Anne Rosser

**Affiliations:** Centre for Neuropsychiatric Genetics and Genomics, Division of Psychological Medicine and Clinical Neurosciences, School of Medicine, Cardiff University, Cardiff, United Kingdom; University Hospital Wales, Cardiff, United Kingdom; Centre for Trials Research, Cardiff University, Cardiff, United Kingdom; UK Dementia Research Institute at Cardiff University, Cardiff, United Kingdom; Cardiff Brain Repair Group, School of Biosciences, Cardiff University, Cardiff, United Kingdom

## Abstract

**Importance:** Antidepressants are the most frequently prescribed medication in Huntington’s disease; this study examines the effect of antidepressants on disease progression.

**Objectives:** Determine the effects on disease progression (composite score, brain atrophy, neurofilament light chain) in Huntington’s disease of (1) psychiatric symptoms and (2) accounting for (1) antidepressant use.

**Design:** Comparison of disease progression between individuals taking antidepressants and non-users, matching for other characteristics via propensity scores.

**Setting:** Global observational cohort studies. Enroll-HD (recruitment 2012-present, annual follow-up) TRACK-HD (recruitment 2008-2013, annual follow-up for 36 months).

**Participants:** Adult with genetically-confirmed repeat expansion for Huntington’s disease not on antidepressants at baseline visit.

**Exposures:** (1) An episode of psychiatric symptoms (depression or anxiety occurring after baseline: problem behaviours assessment score >4 or hospital anxiety and depression score >7) (2) antidepressant use (WHO ATC code N06A) following a new episode of psychiatric symptoms.

**Main Outcome(s) and Measure(s):** The clinical outcome measure was the annual change in composite score of disease progression (two cognitive task scores, a functional score and the motor score, derived from the unified HD rating scale). Biomarker outcomes were change from baseline to 3-year follow-up in a) neurofilament light chain b) brain atrophy in caudate, putamen, whole brain, gray matter, white matter and ventricles.

**Results:** Psychiatric symptoms (3131/6166 Enroll-HD: respective age&sex 47.93(13.81) 56% female,47.37(14.5) 50% female; 115/165 TRACK-HD: age&sex 47.1(9.63) 55% female,48.82(11.6) 48% female) were associated with faster disease progression, increasing composite score decline from 0.38 to 0.58/year (95%CI 0.15,0.25;p=1.2×10-14) and increased rise in neurofilament light chain by 5.3pg/ml (95%CI 1.58,9.024;p=0.007). Antidepressant naive HD participants with new depression or anxiety who started antidepressants (Enroll-HD 194/1877; respective age&sex 52.13(11.77) 57% female, 49.91(13.62) 55% female, TRACK-HD 6/55; respective age&sex 46(3.02) 67% female, 47.22(10.17) 35% female) had reduced composite score decline from 0.89 to 0.53/yr (95%CI 0.13-0.6; p=0.002); a smaller increase of neurofilament light chain by 6.77pg/ml (95%CI 1.8-11.6; p=0.011) and reduced brain atrophy across multiple regions (caudate, putamen, whole brain, gray matter).

**Conclusions and Relevance:** Antidepressant use is associated with slower disease progression in HD on both clinical measures and biomarkers of disease progression. This may have relevance to other neurodegenerative diseases.

---

Psychiatric symptoms are very common in Huntington’s disease (HD); a progressive neurodegenerative disorder focussed on cortico-striatal networks, that is caused by a CAG repeat expansion in the *Huntingtin* gene^1^. The commonest psychiatric symptoms in HD are depression, irritability and apathy^2^. These symptoms have a significantly higher impact on function and quality of life in HD than motor impairments do^3^. Consequently, antidepressants and other psychoactive medications are very frequently prescribed to patients with HD^4^.

The evidence base supporting the use of psychoactive medication in HD is limited: no randomised controlled trials of antidepressants for depression as a primary outcome in HD have been performed, or are listed on trial registries. Furthermore, evidence from other neurodegenerative diseases such as Alzheimer’s disease suggest that antidepressants are ineffective for depression^5^; and psychoactive medication is associated with higher all cause mortality in people with dementia^6^. In HD specifically, recent work has suggested faster disease progression in patients with HD treated with antidepressants^7^.

However, observational studies showing excess mortality and/or faster disease progression in neurodegenerative diseases have not addressed confounding by indication. Prescription of antidepressants and other psychoactive medications may reflect more severe clinical symptoms (and potentially more severe neurodegeneration) necessitating more aggressive symptomatic treatment. Further to this, previous studies showing worsening clinical measures of disease progression in HD, have not included biomarkers of disease progression.

This work addresses the effects of antidepressants on disease progression by using data drawn from two large observational studies of HD: ENROLL-HD^8^ and TRACK-HD^9^. ENROLL-HD is a large international observational study of HD in which participants at all disease stages are followed longitudinally using a range of clinical assessments and self-report scales. TRACK-HD is a smaller longitudinal study of premanifest and early manifest gene-positive individuals with associated collection of blood and CSF for biomarker analysis.

This study had the following objectives:

1. Determine the effect of incident psychiatric symptoms on a) clinical disease progression and b) imaging and CSF biomarkers of disease progression.
2. In antidepressant-naive participants with incident psychiatric symptoms, compare a) clinical disease progression and b) imaging and CSF biomarkers of disease progression between those starting an antidepressant versus no treatment.

## Methods

For this study we used data from two observational studies, ENROLL-HD^8^ and TRACK-HD^9^. ENROLL-HD is an ongoing, worldwide observational study of people with HD and familial controls. Information is collected yearly with formal assessments of cognitive, psychiatric and motor symptoms, in addition to information on co-morbidities, medication, substance misuse and mental health (suicide attempts, admission to inpatient mental health facility admission). The most recent release (periodic data set 6 – PDS6) included data from 25 550 individuals, with the earliest data being collected in 2012(mean visit 3.06, visit range 1-15; including unscheduled visits). TRACK-HD and TRACK-ON included 366 individuals with HD and familial controls. Participants in TRACK-HD also had yearly clinical assessments (at 0,12,24 and 36 months), with a wider range of cognitive and motor assessments than ENROLL-HD. Participants in TRACK-HD also had plasma neurofilament light chain (NfL) collected at study entry and final visit (36 months following baseline); in addition to yearly volumetric MRI scans. Both TRACK-HD and ENROLL-HD adhered to the declarations of Helsinki, and all recruited patients were formally consented. Both studies received institutional approval from UK research ethics committees.

In both datasets, we included all patients aged 18 or over, with a confirmed genetic diagnosis of HD. We excluded any HD participants already receiving antidepressants at baseline study visit to compare the trajectories of disease progression after starting an antidepressant versus not starting one.

The commonest indications for antidepressant use were anxiety and depression, accounting for over 80% listed indications (Table 1, eMethods). We therefore selected incident episodes of depression or anxiety occurring after study entry as the indication for antidepressant prescription.

Psychiatric symptoms are measured by the Problem Behaviours Assessment(short form^8^ - PBAs) in both TRACK-HD and ENROLL-HD, and the Hospital Anxiety and Depression Scale(HADS^8^) in ENROLL-HD. The PBAs has multiple subscales measuring depressed mood, anxiety and 9 other psychiatric symptoms common in HD. Each symptom is scored from 0-4 on both severity and frequency, the product of severity and frequency ranges from 0-16. The HADS depression score (0-21) and anxiety score (0-21) are self report scores, with higher scores indicating more severe symptoms. We determined an episode of anxiety or depression as the relevant PBAs subscale>4, or relevant HADS subscale >7.

As clinical outcome variables in ENROLL-HD, we used the most reliable disease progression measure: composite disease score^10^. This comprises a combination of two cognitive task scores (Stroop word reading task – SWRT, and symbol digit modality task – SDMT^8^), the functional scale (total functional capacity – TFC) and the motor score from the unified HD rating scale ^11^ (UHDRS motor score). The composite score becomes more negative with increasing disease progression. We included the composite score components as secondary outcomes: the TFC, SWRT and SDMT all decline with disease progression, whilst the UHDRS motor score increases with disease progression.

As outcome variables in TRACK-HD, we used change from baseline in plasma neurofilament light chain (plasma NfL), and change from baseline in volumetric measures of brain atrophy (as a percentage of intracranial volume, or the boundary shift integral) as described in Tabrizi et al ^9^: whole brain volume, grey matter, white matter, putamen, caudate.

### Statistical Approach

All analyses were conducted using R, an open source statistical analysis package, using the svy2lme, lme4, Sensemakr and Twang packages. The Sidak correction was used to account for multiple comparisons in secondary clinical outcomes, and imaging outcomes.

To determine the effect of anxiety and depression on clinical disease progression, we constructed a linear mixed model, with an outcome of the composite disease score, and included baseline composite disease score, age, sex, and CAG length as co-variates. The independent variable was presence or absence of a new incident episode of anxiety/depression in ENROLL-HD after baseline; we included subject as a random effect on intercept and slope.

To determine the effect of depression and anxiety on biomarkers of progression in TRACK-HD, we constructed a linear model, with an outcome of change from baseline to final follow-up in 1) NfL and 2) brain volume (whole brain volume, caudate, putamen, grey matter, white matter), and included baseline NfL level or relevant brain volume in the relevant model, as well as age, sex, and CAG length as co-variates. The independent variable was presence or absence of an episode anxiety/depression before final follow-up.

We used propensity scoring to determine the effect of antidepressants on disease progression. To avoid confounding by indication (antidepressants are most commonly prescribed to patients with depression and anxiety, who may have worse disease progression), we selected antidepressant-naive patients from ENROLL-HD with incident episodes (i.e. previously non-depressed, non-anxious participants who experienced a de novo episode of depression or anxiety at any point in ENROLL-HD), throughout the study and compared subsequent disease progression in those who started an antidepressant before the next follow-up, with participants who did not. In TRACK-HD, we selected antidepressant naive patients with anxiety or depression at baseline or first follow-up, and compared 1) patients starting an antidepressant before next follow-up to 2) those not starting antidepressants; on changes in imaging and fluid biomarkers of disease progression. We included age; sex; total number of antidepressant changes; baseline scores for PBAs depression, PBAs suicidal ideation, PBAs anxiety, PBAs irritability; concurrent other psychoactive medication use (anti-dopaminergic medication, mood stabilisers, benzodiazapines); significant mental health event (suicide or psychiatric hospital admission); multiple comorbidities(>5); and baseline composite score in the propensity scoring model. All variables were included in the outcome model in addition to the inverse propensity score in a doubly robust process as described in Funk et al^12^. We used the Sensemakr and tipr packages to test for unmeasured confounding, and the nnet and simputation packages to compare missing data between treatment groups and impute missing data.

## Results

### Association Between Psychiatric Symptoms and Disease Progression

In ENROLL-HD, 3131/5996 antidepressant-naive participants with HD who were free of psychiatric symptoms at baseline experienced an episode of depression or anxiety during the study. The group with psychiatric symptoms were older, more likely to be female, with more advanced disease and longer CAG length (eTable 1).

The linear mixed model of the effect of psychiatric symptoms (episode of anxiety or depression) on the composite score showed that patients with psychiatric symptoms progressed faster than those without. The composite score deteriorated by 0.38/year across all participants. The presence of psychiatric symptoms accelerated this deterioration by 0.20 (95%CI 0.15,0.25; p=1.2×10^-14^)(Figure 1a, eTable 2a). In keeping with the effects of psychiatric symptoms on the composite score, psychiatric symptoms affected the individual composite score components in a similar fashion (eTables 2b-e). We found more rapid disease progression in participants experiencing psychiatric symptoms compared to those without, with a similar effect size across subcomponents of the composite score (30-40% more rapid disease progression/year), all effects were highly significant.

**Figure 1:**
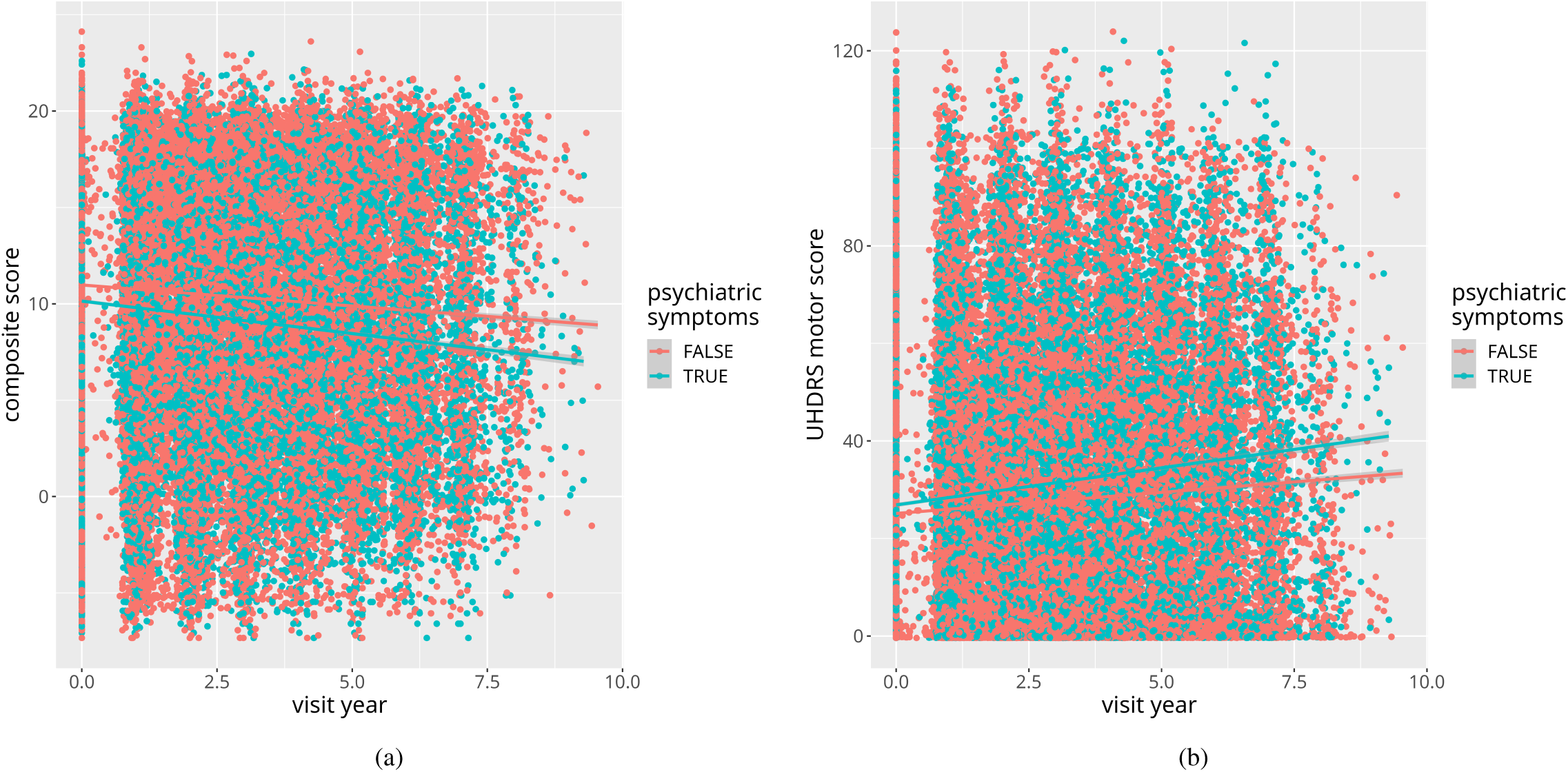
Psychiatric Symptoms Effect on a) Composite Score Progression and b) UHDRS Motor Score Progression.

In TRACK-HD, 115/165 antidepressant-naive participants with HD experienced an episode of psychiatric symptoms before the final visit. Participants with psychiatric symptoms were younger than those without, with a higher proportion of females, more advanced disease and longer CAG lengths (eTable 3). No differences between those with an episode of psychiatric symptoms compared to those without were seen on any measure of brain atrophy after correction for multiple comparisons. However, psychiatric symptoms were associated with larger change from baseline in NfL (5.3pg/ml; 95%CI 1.58,9.024; p=0.007), consistent with more rapid disease progression in this group (eTable 4).

### Associations Between Antidepressant Use and Disease Progression

We selected patients with a new episode of depression or anxiety (i.e. any participant previously free of psychiatric symptoms, developing an incident episode at any point in ENROLL-HD) and compared subsequent disease progression on the composite score between the group receiving antidepressants before the next follow-up visit, to those not treated. In ENROLL-HD, 1877 antidepressant-naive participants (who had been previously free of depression and anxiety) experienced a new episode of psychiatric symptoms: 194 were treated with antidepressants (10.33%). The treated group were significantly different from the untreated across a number of variables: the treated group were older, with more advanced disease, higher scores on all psychiatric variables (depression, suicidality, anxiety, irritability), more psychoactive drug use, more frequent antidepressant treatments across the study, more frequent mental health events, but shorter CAG lengths (eTable 5).

Participants treated with antidepressants had slower decline in the composite score compared to the untreated group (Figure 2a, eTable 6a); per year, the composite score declined by 0.89 in this group, antidepressants lessened this decline by 0.36 (95%CI 0.13,0.6;p=0.002). Of the subcomponents of the composite score (TFC, SDMT, SWRT, motor score) only motor score progression was significantly affected by antidepressant use (Figure 2b, eTable 6b) after accounting for multiple comparisons. Participants receiving an antidepressant had slower increase in motor score than untreated patients: per year, motor score increased by 4.5, antidepressant use reduced this by 1.55(95%CI 0.55,2.55;p=0.0022). Both the TFC and SDMT showed similar effects, but did not meet significance after Sidak correction (p=0.013).

**Figure 2:**
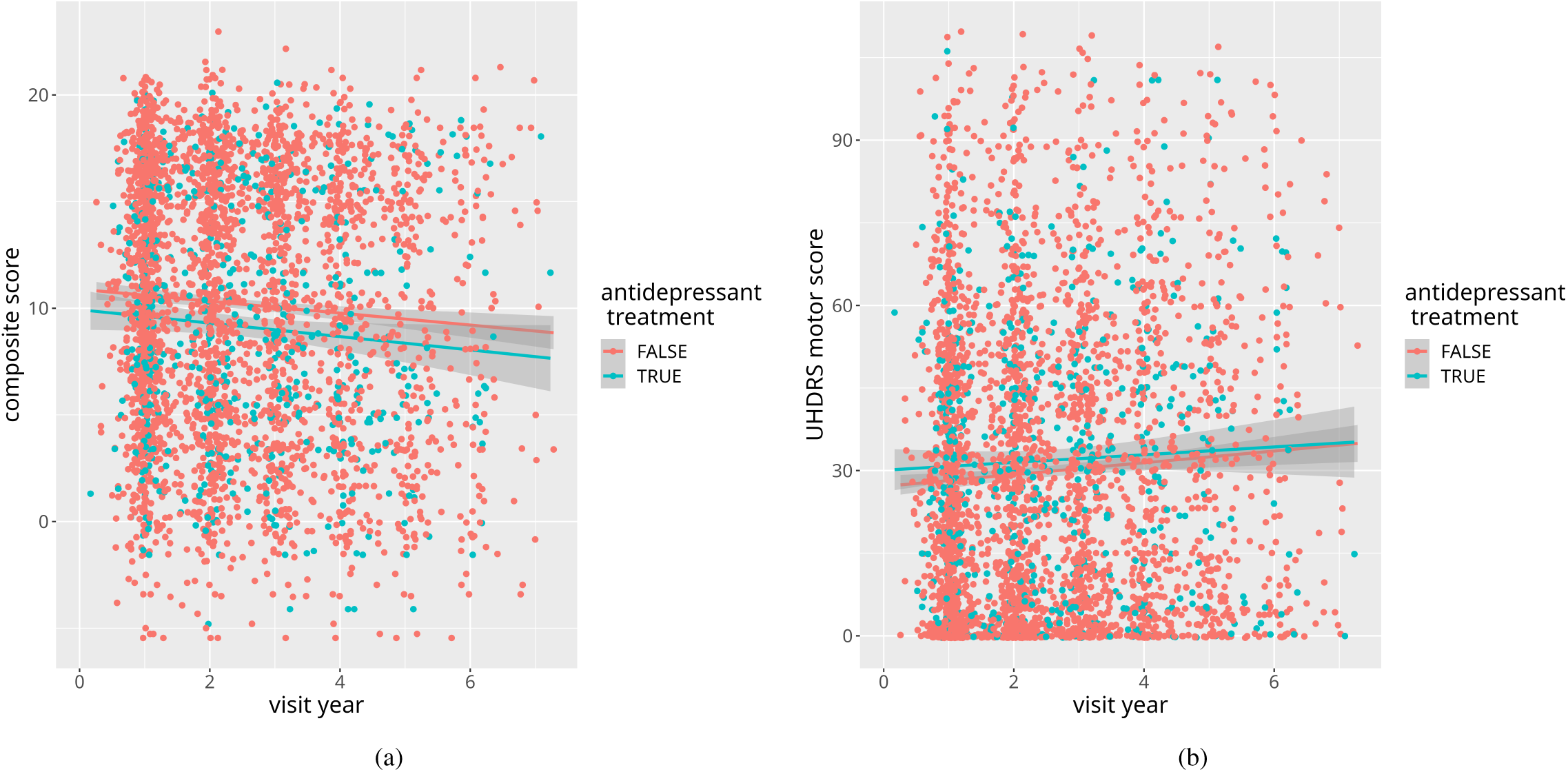
Effect of Antidepressants on a) Composite Score Progression and b) UHDRS Motor Score Progression.

In TRACK-HD, 55 antidepressant-naive patients developed a new episode of depression or anxiety (at baseline or first follow-up) of whom 6 were treated with antidepressants before the next study visit. The treated group were more likely to be male, had more advanced disease, higher psychiatric scores (depression, anxiety, irritability, suicidality), more mental health events, more psychoactive drug use and more frequent antidepressant treatments as the study progressed (eTable 7).

Antidepressant-naive participants with a new episode of anxiety or depression (at first or second study visit) treated with antidepressants had a smaller change from baseline in plasma NfL (group difference -6.77pg/ml;95%CI 1.8,11.6; p=0.011)(Figure 3a, eTable 8a). This is consistent with slower disease progression in HD driven by antidepressant use.

**Figure 3:**
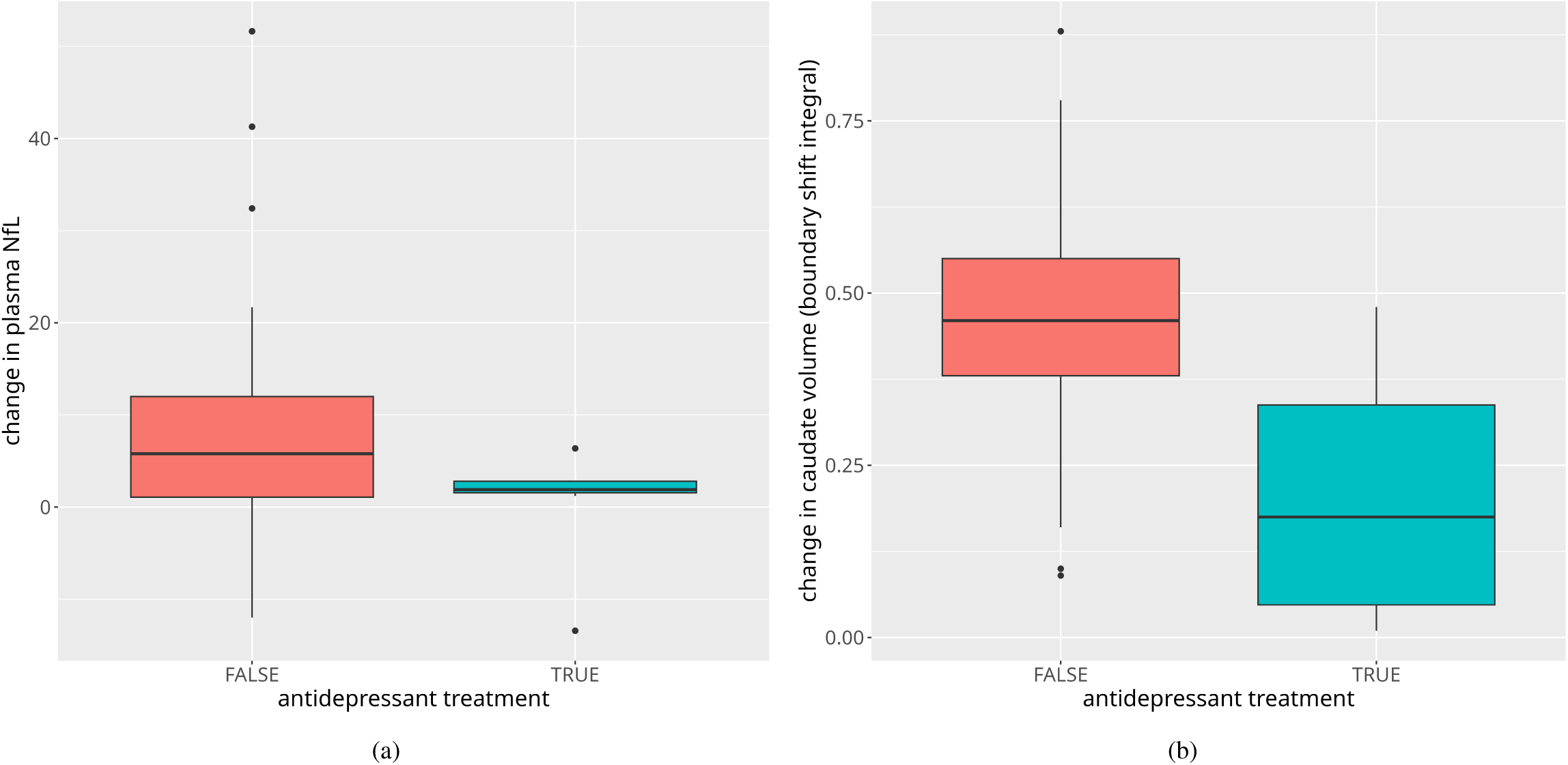
Effect of Antidepressants on a) Change in Plasma NfL and b) Caudate Atrophy.

We found similar effects of antidepressants on atrophy rates in all imaging variables except ventricular volume (p=0.83)(Figures 3b,4a&4b, eTables 8b-e). Change from baseline in caudate volume was smaller in the group treated with antidepressants compared to the untreated group(group difference in boundary shift integral; -0.23; 95%CI 0.18,0.28; p=1.2×10^-12^), as were whole brain volume(group difference in boundary shift integral; -11; 95%CI 5.71,16.29; p=0.00018), putamen (group difference volume loss from baseline as percentage of intracranial volume: -0.0001;95%CI 0.00013,0.000073; p=2.4×10^-10^) and grey matter (group difference in volume loss from baseline as percentage of intracranial volume: -13; 95%CI 4.57,21.43 p=0.0043). White matter volume atrophy was also reduced in patients on antidepressants (p=0.012), but this did not satisfy the Sidak correction for multiple comparisons in imaging measures (0.0073).

**Figure 4:**
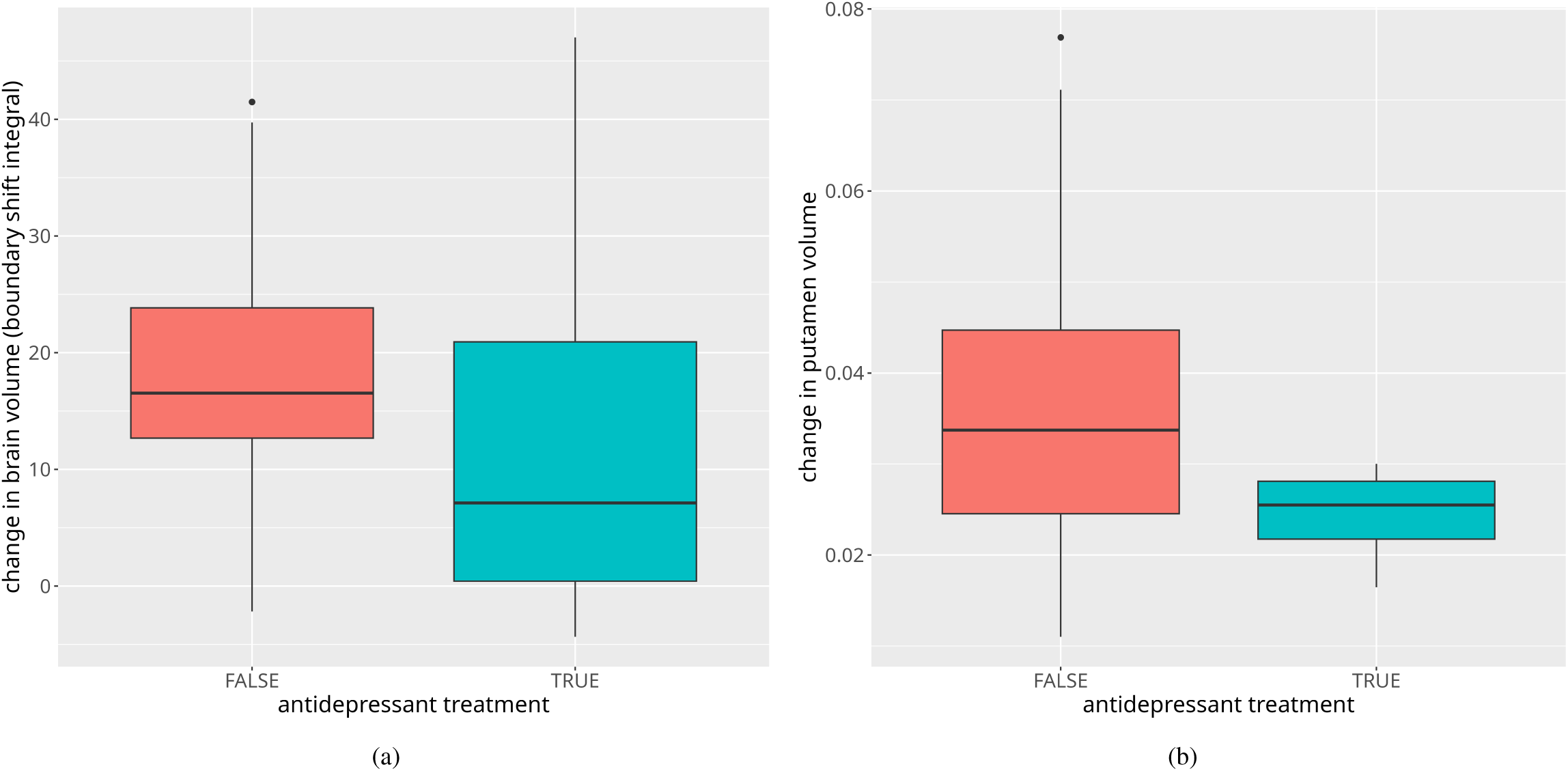
Effect of Antidepressants on a) Whole Brain Atrophy and b) Putamen Atrophy.

A sensitivity analysis in the ENROLL-HD dataset showed that an unobserved confounder accounting for more than 4.98% of the residual variation in composite score would eliminate the effect of the interaction between antidepressant treatment and time on composite score. This is less than the effect of baseline composite score and time, but more than comorbidities, psychiatric scores, CAG length, age, sex, history of addiction, antidepressant burden and previous mental health events (eTable 9). Missing data did not differ between treatment groups (eMethods), sequential hotdeck imputation of missing data did not change the association between antidepressant treatment and composite score, UHDRS motor score, NfL, caudate volume or whole brain volume(eTables 10a-e), although putamen and grey matter volume were no longer significant.

## Discussion

In this work, we have shown that in HD participants with new episodes of depression or anxiety, starting an antidepressant is associated with slower disease progression on both clinical measures and imaging and plasma biomarkers of disease progression. As psychiatric symptoms are common in neurodegenerative diseases more widely, the effect of antidepressants on disease progression in these disorders merits further study. Significantly, we have also shown that in HD, psychiatric symptoms are associated with faster disease progression: both on clinical and biomarker measures of disease progression.

Previous work in the ENROLL-HD dataset, had suggested more rapid disease progression in patients treated with antidepressants^7^. However, the model in this study did not include relevant mental health variables, and did not select participants experiencing incident mental health problems introducing confounding by indication. More recently a propensity score approach was used in motor-symptomatic participants in ENROLL-HD^13^. This study included depression and anxiety scores, CAG length, composite score components in the model, with a primary outcome of depression score at first follow up visit after initiation. They found 86 new users of antidepressants, but did not find differences at first follow-up on depression or any measure of disease progression. We looked at disease progression over time, included a substantially larger N and found an effect on imaging and fluid biomarkers of disease progression. A number of studies have also noted an association between antidepressant use and the onset of cognitive decline or diagnosis of dementia^14,15^, though these studies were not able to account for baseline psychiatric symptom severity. More recent observational work in Alzheimer’s has shown some effect of tricyclic antidepressants (but not other antidepressants) in worsening cognitive decline, but no effect on imaging biomarkers of disease progression^16^. In Parkinson’s disease, antidepressants were initially linked with a higher risk of disease onset in an observational study^17^, although animal work in PD models has shown beneficial effects of antidepressant treatment on alpha synuclein deposition^18^, and a trial of antidepressants for depression in Parkinson’s disease has included disease progression as a secondary outcome.

To date, no clinical trial of antidepressants in HD (for any indication) has included more than 30 participants in each arm and the follow-up has been limited to several months. Randomised controlled trials (summarised in Zadegan et al^19^) of fluoxetine (primary outcome – function, measured by the TFC), citalopram (primary outcome – cognition), bupropion (primary outcome – apathy) and a novel agent PNU-96391A (primary outcome – safety and tolerability) all had follow-up periods shorter than 6 months and 20 or fewer participants in each arm. None showed any benefit on established clinical measures of disease progression, but previous work in HD has shown the need for a substantially larger sample size and longer follow-up^10^.

The association between psychiatric symptoms and disease progression in HD suggests that there may be an additional mechanism leading to disease progression other than that driven by CAG repeat length, represented phenotypically by psychiatric symptomatology. As psychiatric symptoms are more frequent in many neurodegenerative diseases compared to the wider population, this raises the intriguing possibility that there is a common mechanism underlying these symptoms across neurodegenerative diseases, potentially opening novel therapeutic avenues. Determining what this process might be is difficult as the mechanism(s) leading to depression are uncertain. Previous work by our group has shown overlap with genetic risk for psychiatric disorders in neurodegenerative disease and the general population^20,21^. Impairments in neurogenesis have been found in patients with depression in the general population, are known to be rescued by antidepressant treatment, and have been found in R6/1 animal models of HD that can be rescued by fluoxetine treatment^22^. Hypothalamo-pituitary axis dysfunction has been found in depression in the general population and HD^23^. Evidence of CNS inflammation has been found in depression^24^, and also in HD - with some data to suggest modification in imaging biomarkers of disease progression with immunomodulatory treatment^25^.

There are a number of limitations in this study. Propensity score matching is not able to account for unobserved confounders, however there is some evidence to suggest that observed and unobserved confounders co-vary^26^. Further to this, a sensitivity analysis suggested that the observed effect would be eliminated by a confounder with an influence larger than all co-variates other than baseline composite score and time– it is difficult to conceive of a confounder of this magnitude, missing from the model. The number of the subjects in the TRACK-HD data is small, however the effects are all in the same direction, satisfy correction for multiple comparisons and there are trend level effects (white matter atrophy) that are in the same direction as the significant effects.

In conclusion, in this work we have shown slower disease progression on clinical, imaging biomarker and fluid biomarkers in HD patients treated with antidepressants compared to untreated patients. This finding may be of wider applicability to other neurodegenerative diseases, and raises the possibility of a second mechanism contributing to disease progression in HD.

## Data Availability

All data produced in the present study are available upon reasonable request to the authors

## Acknowledgements and Conflict of Interest

Financial: Dr Drew has received financial honoraria for reviewing clinical trial protocols as part of the Enroll-HD clinical trials committee, Professor Holmans is paid an honorarium of $2000 per annum as part of the scientific review committee for Enroll-HD.

Leadership: Dr McLauchlan is the chair of the SBAC committee of the EHDN a grant awarding body, Professor Holmans is a member of the SBAC. Professor Rosser is the previous President of the EHDN.

## Funding

ENROLL-HD and TRACK-HD were funded by CHDI Foundation. Dr McLauchlan is funded by a Health and Care Research Wales Research Time award. Cheney Drew is funded by Health Care Research Wales, Michael J Fox Foundation and Jacques and Gloria Gossweiler Foundation. All other authors are funded by Cardiff University.

## Author Roles

Duncan J McLauchlan: conceptualisation, data curation, formal analysis, funding acquisition, methodology, writing - original draft

Cheney JG Drew: conceptualisation, writing - reviewing and editing

Peter Holmans: investigation, methodology, formal analysis, writing - reviewing and editing

Anne Rosser: methodology, supervision, writing - reviewing and editing

## Notes

### Author Declarations

NHS Research Ethics Committee of Wales gave approval of this work (13/WA/0192)

