## supplemental file 1 for "Antidepressants and Slower Disease Progression in Huntington’s Disease"

### SUPPLEMENTARY DATA

**Electronic Methods**

First, we determined the frequency of antidepressant use in HD: antidepressants were the most frequently prescribed medication to patients with HD in the ENROLL-HD dataset, accounting for over 15% of all drugs prescribed. We categorised all listed indications for antidepressants as ‘depression’, ‘anxiety’, ‘irritability and aggression’, ‘apathy’, ‘sleep’, ‘psychosis’, ‘other psychiatric symptom’, ‘motor symptom of HD’, ‘pain’, ‘systemic symptom’ or ‘uncertain’ as outlined below.

Depression

”Adjustment disorder”,”Adjustment disorder with depressed mood”,”Adjustment dis- order with mixed anxiety and depressed mood”, ”Affect lability”,”Affective disor- der”,”Agitated depression”, ”Antidepressant therapy”,”Depressed mood”,”Depression”, ”Depressive symptom”, ”Dysthymic disorder”, ”Grief reaction”, ”Major depression”, ”Mood altered”,”Perinatal depression”, ”Mood disorder due to a general medical con- dition”,”Negative thoughts”, ”Postpartum depression”,”Seasonal affective disorder”, ”Suicidal ideation”,”Suicide attempt”,”Suicidal behaviour”, ”Tearfulness”, ”Negativism”, ”Persistent depressive disorder”.

Anxiety

”Acute stress disorder”,”Agoraphobia”,”Anxiety”, ”Anxiety disorder”,”Anxiolytic ther- apy”, ”Anticipatory anxiety”, ”Claustrophobia”,”Phobia of flying”,”Mixed anxiety and depressive disorder”, ”Compulsions”, ”Generalised anxiety disorder”, ”Nervousness”, ”Neurosis”, ”Obsessive thoughts”, ”Compulsions”, ”Obsessive-compulsive disorder”, ”Obsessive-compulsive personality disorder”, ”Panic attack”, ”Panic disorder”, ”Pho- bia”, ”Post-traumatic stress disorder”, ”Social phobia”, ”Stress”, ”Tension”, ”Obses- sive rumination”,”Social anxiety disorder”.

Apathy

”Apathy”,”Lethargy”, ”Fatigue”

Irritability

”Aggression”,”Anger”,”Irritability”, ”Anger”,”Conduct disorder”, ”Irritability postvac- cinal”,”Intermittent explosive disorder” , ”Mood swings”, ”Impatience”, ”Affect labil- ity”

Psychosis

”Delusion”,”Delusional disorder, unspecified type”,”Hallucination”, ”Hallucination vi- sual”,”Jealous delusion”, ”Psychotic disorder”, ”Schizoaffective disorder”, ”Delusions, mixed”, ”Schizoaffective disorder depressive type”, ”Schizophrenia”, ”Schizophrenia paranoid type”,”Schizophrenia, paranoid type”, ”Acute psychosis”, ”Paranoia”,”Somatic Delusion”, ”Psychotic behaviour”, ”Hallucination auditory”, ”Psychotic disorder due to a general medical condition”.

Sleep

”Circadian rhythm sleep disorder”,”Initial insomnia”,”Insomnia”, ”Insomnia related to another medical condition”, ”Middle Insomnia”, ”Narcolepsy”, ”Poor quality sleep”, ”Sedation”, ”Sedative therapy”, ”Sleep disorder”, ”Parasomnia”,”Sleep disorder due to general medical condition, insomnia type”, ”Sleep phase rhythm disturbance”, ”Sleep disorder”, ”Sleep disorder due to a general medical condition, insomnia type”, ”Sleep terror”,”Somnolence”,”Rapid eye movement sleep abnormal”,”Somnolence”,”Rapid eye movements sleep abnormal”.

Other Psychiatric

”Abnormal behaviour”,”Agitation”,”Akathisia”,”Amnesia”,”Abstains from alcohol”, ”Al- cohol abuse”, ”Alcohol withdrawal syndrome”, ”Alcoholism”,”Attention deficit/hyperactivity disorder”, ”Attention deficit hyperactivity disorder”,”Binge eating”,”Bipolar disorder”, ”Bipolar I disorder”,”Bipolar II disorder”, ”Amnesia”,”Autism spectrum disorder”, ”Behaviour disorder”,”Borderline personality disorder”, ”Bradyphrenia”,”Burnout syn- drome”, ”Cyclothymic disorder”,”Confusional state”, ”Cognitive disorder”, ”Conver-

sion disorder”,”Dementia”, ”Drug dependence”,”Depressed level of consciousness”, ”Delirium”,”Detoxification”, ”Disinhibition”, ”Disorientation”, ”Disturbance in social behaviour”,”Disturbance in attention”,”Drooling”, ”Drug withdrawal syndrome”,”Eating disorder”, ”Drug abuse” ,”Emotional distress”,”Emotional disorder”,”Euphoric mood”, ”Eructation”, ”Fear”,”Fear of disease”, ”Feeling of relaxation”, ”Gambling”,”Hyperphagia”, ”Hyperventilation”, ”Impulse-control disorder”, ”Impulsive behaviour”,”Impulse-control disorder”, ”Kleptomania”,”Libido disorder”, ”Loss of libido”,”Memory impairment”, ”Mental disorder”, ”Mental impairment”, ”Mood swings”, ”Mild mental retardation”,”Mania”, ”Malaise”, ”Hypersexuality”, ”Middle insomnia”, ”Nicotine dependence”, ”Nightmare”, ”Paranoid personality disorder”, ”Perseveration”, ”Personality change”,”Personality change due to a general medical condition”, ”Personality disorder”,”Polydipsia psy- chogenic”, ”Psychomotor hyperactivity”,”Psychiatric symptom”, ”Psychosomatic dis-

ease”, ”Relaxation therapy”, ”Smoking cessation therapy”,”Supplementation therapy”, ”Screaming”, ”Schizotypal personality disorder”, ”Somatisation disorder”, ”Somato- form disorder”, ”Thinking abnormal”, ”Trichotillomania”,”Tachyphrenia”, ”Tobacco abuse”, ”Tobacco user”.

Motor Symptom of HD

”Bradykinesia”,”Balance disorder”,”Cerebellar syndrome”,”Clumsiness”,”Chorea”, ”Dys- tonia”,”Dyskinesia”, ”Hypertonia” ,”Hyperkinesia”,”Movement disorder”, ”Muscle re- laxant therapy”, ”Extrapyramidal disorder”,”Hypotonia”,”Myoclonus”, ”Oromandibu- lar dystonia”,”Parkinsonism”,”Muscle spasticity” ,”Muscle rigidity”, ”Periodic limb movement disorder”, ”Restless legs syndrome”, ”Torticollis” ,”Tremor”,”Tardive dysk- inesia”, ”Tic”.

Pain

”Abdominal pain”,”Abdominal pain upper”,”Analgesic therapy”,”Abdominal tender- ness”,”Allodynia”,”Anaesthesia”,”Arthralgia”,”Arthritis”, ”Back pain”,”Chest pain”, ”Burn- ing sensation”,”Complex regional pain syndrome”,”Facial Neuralgia”,”Facial pain”, ”Fibromyalgia”, ”Headache”, ”Hernia pain”,”Epicondylitis”, ”Gastrointestinal pain”, ”Arthropathy”, ”Meralgia paraesthetica”, ”Migraine”, ”Migraine prophylaxis”, ”Mus- culoskeletal pain”, ”Neck pain”,”Myalgia”, ”Neuralgia”, ”Occipital neuralgia”, ”Pain”, ”Pain in extremity”, ”Pain management”, ”Procedural pain”,”Oral discomfort”, ”Post herpetic neuralgia”, ”Psychogenic pain disorder”, ”Radicular pain”,”Rib fracture”,”Sciatica”,

”Tension headache”, ”Trigeminal neuralgia”, ”Vulvovaginal pain”.

Systemic Illness

”Abnormal loss of weight”,”Ankylosing spondylitis”, ”Arthritis”,”Antiallergic ther- apy”,”Autoimmune thyroiditis”,”Asthma”, ”Back injury”,”Bladder disorder”,”Bladder irritation”, ”Bladder spasm”,”Brain neoplasm”, ”Bruxism”,”Brachial plexus injury”,”Carpal tunnel syndrome”, ”Abdominal distension”,”Chills”, ”Chronic fatigue syndrome”,”Cough”, ”Corneal dystrophy”,”Craniocerebral injury”, ”Crohn’s disease”,”Cystitis interstitial”, ”Convulsion”,”Convulsion prophylaxis”,”Cystitis”,”Cyclic vomiting syndrome”,”Dysuria”, ”Dyslipidaemia”, ”Decreased appetite”, ”Dermatitis”,”Dermatitis atopic”,”Diabetic neu- ropathy”,”Dizziness”, ”Duodenal ulcer”, ”Dyspepsia”, ”Dysaesthesia”, ”Dysphagia”,”Dyspnoea”, ”Enuresis”, ”Essential hypertension”,”Ear infection”, ”Eczema”,”Endometriosis”,”Epilepsy”, ”Fatigue”, ”Fall”, ”Gastrooesophageal reflux disease”,”Gastrointestinal reflux disease”, ”Gastrointestinal disorder”, ”Glaucoma”,”Haemorrhoids”, ”Gastritis prophylaxis”,”Head injury”,”Herpes zoster”, ”Hot flush”,”HIV peripheral neuropathy”, ”HIV infection”, ”Hypertension”, ”Hypermobility syndrome”, ”Hyperhidrosis”,”Hypertonic bladder”,”Irritable bowel syndrome”,”Increased appetite”,”Incontinence”,”Intracranial pressure increased”,”Impaired gastric emptying”, ”In vitro fertilisation”,”Intervertebral disc disorder”,”Intervertebral

disc degeneration”,”Intervertebral disc protrusion”,”Hiccups”,”Hiatus Hernia”, ”Joint injury”,”Limb operation”,”Lung neoplasm malignant”,”Macular oedema”,”Meniere’s disease”, ”Menopausal symptoms”, ”Menopause”, ”Menstrual disorder”,”Multiple scle- rosis”, ”Mitral valve prolapse”,”Motion sickness”,”Muscle contracture”,”Muscle relax- ant therapy”,”Microvascular coronary artery disease” ,”Peripheral swelling”, ”Mononeu-

ropathy”,”Muscle spasms”,”Musculoskeletal stiffness”,”Nausea”,”Nerve compression”,”Nerve injury”,”Neuritis”,”Neuropathy peripheral”, ”Oedema peripheral”, ”Obesity”, ”Osteoarthri- tis”,”Overweight”, ”Otitis media chronic”, ”Paraesthesia”, ”Periarthritis”,”Partial seizures”, ”Pruritis”,”Psoriasis”, ”Psoriatic arthropathy”, ”Paraesthesiae oral”, ”Prophylaxis against gastrointestinal ulcer”, ”Polyneuropathy”, ”Petit mal epilepsy”, ”Post-traumatic epilepsy”, ”Post viral fatigue syndrome”, ”Premenstrual syndrome”,”Rhinitis allergic”,”Regurgitation”, ”Salivary hypersecretion”, ”Pollakiuria”,”Photopsia”,”Radiculopathy”,”Scoliosis”, ”Sim-

ple partial seizures”, ”Syncope”, ”Seasonal allergy”, ”Sensory disturbance”, ”Sinusi- tis”, ”Sleep apnoea syndrome”, ”Seizure anoxic”, ”Sexual dysfunction”, ”Sjogrens syndrome”,”Spinal osteoarthritis”,”Spinal column injury”,”Spinal column stenosis”, ”Som- nambulism”, ”Stress urinary incontinence”,”Upper airway obstruction”,”Upper respi- ratory tract infection”, ”Surgery”,”Urinary incontinence”, ”Urge incontinence” , ”Uri- nary tract infection”, ”Urticaria”,”Vascular occlusion” ,”Vertigo”,”Vomiting”,”Seizure”, ”Somatic symptom disorder”, ”Vomiting in pregnancy”,”Temporal lobe epilepsy”, ”Tem-

poromandibular joint syndrome”,”Tonic clonic movements”,”Tinnitus”, ”Underweight”,”Weight control”.

Unclear

”NA”,”Huntington’s disease”,”Hypersensitivity”, ”MISSING”,”NOTAPPL”, ”Off la- bel use”, ”Product used for unknown indication”, ”Prophylaxis”, ”Restlessness”, ”Pal- liative care”, ”Prophylaxis”,”Nervous system disorder”,”Neurological symptom”,”Motor dysfunction”.

### Missing Data

The final dataset included missing rates between 16% and 26% for clinical outcome variables after elimination of implausible outliers (i.e. results lying outside the range of the task or scale):

Composite - 26.9%

Stroop word reading test - 22.2% Symbol digit modality test - 25.5% UHDRS motor score - 17.2%

UHDRS total functional capacity - 16.3%

Missingness was lower in the antidepressant treated group (24.29% vs 27.34%). Using a multinomial model from the nnet R package, we found that missingness was not significantly associated with treatment group (AIC 6264, antidepressant treatment: estimate -0.16, p=0.076). We subsequently constructed a linear mixed model, with random intercept and slope, to determine the effect of missingess on composite score progression (Table S9). This showed that participants with missing values had more severe progression over time. As missing rates were higher in the control group, this suggests that, if anything, the effect of antidepressant treatment on clinical progression is likely to be underestimated. Similarly, in the TRACK-HD data, there was no asso- ciation between missingness and treatment group, whilst the only association between disease severity biomarkers and missingness was with smaller baseline caudate vol- ume.

Effect of Missingness on Composite Score

|  | Estimate | Std. Error | t value | p value |
| --- | --- | --- | --- | --- |
| (Intercept) | 11 | 0.17 | 1*.*7 *×* 10^3^ | *<*2x10*^−^*^16^ |
| missing data | -1.6 | 0.28 | 1*.*7 *×* 10^3^ | 1*.*2 *×* 10*^−^*^8^ |
| visit year | -0.6 | 0.032 | 6*.*9 *×* 10^2^ | *<*2x10*^−^*^16^ |
| missing data: visit year | -0.26 | 0.05 | 6*.*9 *×* 10^2^ | 2*.*6 *×* 10*^−^*^7^ |

eTable 1: Demographics Psychiatric Symptoms ENROLL-HD

Psychiatric Symptoms Present	Psychiatric Symptoms Absent

| age | 47.93 sd 13.81 | 47.37 sd 14.5 |
| --- | --- | --- |
| sex | 55.55% F | 50.25% F |
| CAG length | 43.23 sd 3.36 | 42.97 sd 3.45 |
| composite score | 12.2 sd5.25 | 13.25 sd 5.1 |

mean and standard deviation (sd) shown

eTable 2a: Psychiatric Symptoms Effect on Composite Score

|  | Estimate | Std. Error | t value | p value |
| --- | --- | --- | --- | --- |
| (Intercept) | 0.9 | 0.2 | 4.5 | 6*.*3 *×* 10*^−^*^6^ |
| psychiatric symptoms | 0.04 | 0.017 | 2.3 | 0.019 |
| visit year | -0.38 | 0.02 | -18 | *<*2 *×* 10*^−^*^16^ |
| age | -0.0042 | 0.00091 | -4.7 | 2*.*8 *×* 10*^−^*^6^ |
| sex (male) | -0.00078 | 0.017 | -0.047 | 0.96 |
| CAG length | -0.016 | 0.0034 | -4.8 | 1*.*9 *×* 10*^−^*^6^ |
| baseline composite score | 1 | 0.0025 | 4 *×* 10^2^ | *<*2 *×* 10*^−^*^16^ |
| psychiatric symptoms: visit year | -0.2 | 0.026 | -7.7 | 1*.*2 *×* 10*^−^*^14^ |

eTable 2b: Psychiatric Symptoms Effect on UHDRS Motor Score

|  | Estimate | Std. Error | t value | p value |
| --- | --- | --- | --- | --- |
| (Intercept) | -5.9 | 0.91 | -6.4 | 1*.*2 *×* 10*^−^*^10^ |
| psychiatric symptoms | -0.21 | 0.095 | -2.2 | 0.028 |
| visit year | 1.7 | 0.092 | 18 | *<*2 *×* 10*^−^*^16^ |
| age | 0.029 | 0.0048 | 6.1 | 1*.*5 *×* 10*^−^*^9^ |
| sex (male) | -0.1 | 0.093 | -1.1 | 0.29 |
| CAG length | 0.11 | 0.018 | 6.3 | 3*.*9 *×* 10*^−^*^10^ |
| baseline UHDRS motor score | 0.98 | 0.0033 | 3 *×* 10^2^ | *<*2 *×* 10*^−^*^16^ |
| psychiatric symptoms: visit year | 0.67 | 0.11 | 5.8 | 7*.*7 *×* 10*^−^*^9^ |

eTable 2c: Psychiatric Symptoms Effect on UHDRS TFC Score

|  | Estimate | Std. Error | t value | p value |
| --- | --- | --- | --- | --- |
| (Intercept) | 1.2 | 0.18 | 6.7 | 1*.*6 *×* 10*^−^*^11^ |
| psychiatric symptoms | 0.0094 | 0.017 | 0.54 | 0.59 |
| visit year | -0.29 | 0.018 | -16 | *<*2 *×* 10*^−^*^16^ |
| age | -0.0049 | 0.00078 | -6.3 | 2*.*7 *×* 10*^−^*^10^ |
| sex (male) | -0.017 | 0.017 | -1 | 0.31 |
| CAG length | -0.019 | 0.003 | -6.3 | 3 *×* 10*^−^*^10^ |
| baseline UHDRS TFC score | 0.99 | 0.0037 | 2*.*7 *×* 10^2^ | *<*2 *×* 10*^−^*^16^ |
| psychiatric symptoms: visit year | -0.13 | 0.023 | -5.6 | 2*.*5 *×* 10*^−^*^8^ |

TFC - total functional capacity

eTable 2d: Psychiatric Symptoms Effect on Symbol Digit Modality Score

|  | Estimate | Std. Error | t value | p value |
| --- | --- | --- | --- | --- |
| (Intercept) | 11 | 0.99 | 11 | *<*2 *×* 10*^−^*^16^ |
| psychiatric symptoms | 0.15 | 0.088 | 1.6 | 0.1 |
| visit year | -0.48 | 0.067 | -7.2 | 9*.*5 *×* 10*^−^*^13^ |
| age | -0.053 | 0.0047 | -11 | *<*2 *×* 10*^−^*^16^ |
| sex (male) | -0.086 | 0.086 | -1 | 0.32 |
| CAG length | -0.17 | 0.017 | -9.9 | *<*2 *×* 10*^−^*^16^ |
| baseline SDMT score | 0.97 | 0.0036 | 2*.*7 *×* 10^2^ | *<*2 *×* 10*^−^*^16^ |
| psychiatric symptoms: visit year | -0.57 | 0.084 | -6.8 | 1*.*1 *×* 10*^−^*^11^ |

SDMT - symbol digit modality score

eTable 2e: Psychiatric Symptoms Effect on Stroop Word Reading Score

|  | Estimate | Std. Error | t value | p value |
| --- | --- | --- | --- | --- |
| (Intercept) | 18 | 1.6 | 11 | *<*2 *×* 10*^−^*^16^ |
| psychiatric symptoms | 0.089 | 0.16 | 0.57 | 0.57 |
| visit year | -1.7 | 0.12 | -14 | *<*2 *×* 10*^−^*^16^ |
| age | -0.074 | 0.0074 | -9.9 | *<*2 *×* 10*^−^*^16^ |
| sex (male) | -0.01 | 0.15 | -0.067 | 0.95 |
| CAG length | -0.26 | 0.028 | -9.3 | *<*2 *×* 10*^−^*^16^ |
| baseline SWRT score | 0.96 | 0.0039 | 2*.*5 *×* 10^2^ | *<*2 *×* 10*^−^*^16^ |
| psychiatric symptoms: visit year | -0.57 | 0.15 | -3.8 | 0.00018 |

SWRT - Stroop word reading score

eTable 3: Demographics Psychiatric Symptoms TRACK-HD

Psychiatric Symptoms Present	Psychiatric Symptoms Absent

| age | 47.1 sd 9.63 | 48.82 sd 11.6 |
| --- | --- | --- |
| sex | 54.78% F | 48% F |
| CAG length | 43.98 sd 2.86 | 43.24 sd 3.31 |
| composite score | 15.44 sd 2.38 | 15.62 sd 2.86 |

mean and standard deviation (sd) shown

eTable 4: Psychiatric Symptoms Effect on Increase in NfL

|  | Estimate | Std. Error | t value | p value |
| --- | --- | --- | --- | --- |
| (Intercept) | 3.4 | 28 | 0.12 | 0.9 |
| psychiatric symptoms | 5.3 | 1.9 | 2.7 | 0.007 |
| baseline NfL | 0.038 | 0.045 | 0.83 | 0.41 |
| age | -0.09 | 0.13 | -0.67 | 0.51 |
| sex | 3.1 | 1.8 | 1.7 | 0.089 |
| CAG length | -0.17 | 0.45 | -0.38 | 0.71 |
| baseline composite score | 0.47 | 0.46 | 1 | 0.31 |

NfL - neurofilament light chain

eTable 5: Demographics Antidepressant Treatment ENROLL-HD

|  | antidepressant treatment | control | unweighted p value |
| --- | --- | --- | --- |
| age | 52.13 sd 11.77 | 49.91 sd 13.62 | *<*0.001 |
| sex (female) | 0.57 sd 0.49 | 0.55 sd 0.50 | 0.25 |
| baseline composite score | 10.77 sd 5.11 | 11.06 sd 5.70 | 0.16 |
| baseline pba depression score | 4.43 sd 3.79 | 3.55 sd 3.50 | *<*0.001 |
| baseline pba irritability score | 3.15 sd 3.25 | 2.83 sd 3.25 | 0.01 |
| baseline pba anxiety score | 5.89 sd 3.97 | 5.50 sd 3.64 | 0.01 |
| baseline pba suicidality score | 0.86 sd 2.37 | 0.34 sd 1.35 | *<*0.001 |
| number of antidepressant | 2.78 sd 1.81 | 0.79 sd 1.29 | *<*0.001 |
| previous mental health event | 0.52 sd 0.50 | 0.32 sd 0.47 | *<*0.001 |
| history of addiction | 0.57 sd 0.49 | 0.55 sd 0.50 | 0.16 |
| psychoactive drug | 0.76 sd 0.43 | 0.53 sd 0.50 | *<*0.001 |
| CAG length | 43.08 sd 2.60 | 43.37 sd 3.35 | 0.01 |
| comorbidities | 0.41 sd 0.49 | 0.32 sd 0.47 | *<*0.001 |

mean and standard deviation (sd) shown

eTable 6a: Antidepressant Effect on Composite Disease Score

|  | Estimate | Std. Error | t value | p value |
| --- | --- | --- | --- | --- |
| (Intercept) | 9.85 | 5.14 | 1.92 | 0.055 |
| antidepressant treatment | -0.77 | 0.34 | -2.2 | 0.023 |
| visit year | -0.89 | 0.094 | -9.49 | *<*2 *×* 10*^−^*^16^ |
| antidepressant treatment: visit year | 0.36 | 0.12 | 3.085 | 0.002 |

eTable 6b: Antidepressant Effect on UHDRS Motor Score

|  | Estimate | Std. Error | t value | p value |
| --- | --- | --- | --- | --- |
| (Intercept) | -47.82 | 29.24 | -1.64 | 0.1 |
| antidepressant treatment | 4.5 | 1.91 | 2.36 | 0.018 |
| visit year | 3.47 | 0.37 | 9.18 | *<* 2x10*^−^*^16^ |
| antidepressant treatment: visit year | -1.55 | 0.51 | -3.064 | 0.0022 |

eTable 7: Demographics Antidepressant Treatment TRACK-HD

|  | antidepressant treatment | control | unweighted p value |
| --- | --- | --- | --- |
| age | 46.00 sd 3.02 | 47.22 sd 10.17 | 0.30 |
| sex (female) | 0.75 sd 0.45 | 0.35 sd 0.48 | *<*0.001 |
| baseline composite score | 15.33 sd 1.81 | 16.10 sd 2.10 | 0.10 |
| baseline pbs depression score | 8.12 sd 5.92 | 3.81 sd 2.65 | *<*0.001 |
| baseline pba irritability score | 6.12 sd 4.49 | 2.77 sd 2.62 | *<*0.001 |
| baseline pba anxiety score | 8.88 sd 5.12 | 6.40 sd 2.86 | 0.05 |
| baseline pba suicidality score | 3.12 sd 5.30 | 0.54 sd 1.34 | 0.05 |
| number of antidepressant | 1.38 sd 0.50 | 0.73 sd 1.15 | *<*0.001 |
| previous mental health event | 0.00 | 0.19 sd 0.40 | *<*0.001 |
| history of addiction | 0.25 sd 0.45 | 0.31 sd 0.47 | 0.59 |
| psychoactive drug | 0.75 sd 0.45 | 0.26 sd 0.44 | *<*0.001 |
| CAG length | 43.12 sd 1.67 | 44.04 sd 2.88 | 0.05 |
| comorbidities | 0.00 | 0.22 sd 0.42 | *<*0.001 |

eTable 8a: Plasma NfL Change from Baseline

|  | Estimate | Std. Error | t value | p value |
| --- | --- | --- | --- | --- |
| (Intercept) | -64 | 47 | -1.3 | 0.18 |
| antidepressant treatment | -6.7 | 2.5 | -2.6 | 0.011 |
| baseline NfL | -0.0021 | 0.14 | -0.015 | 0.99 |

NfL - neurofilament light chain

eTable 8b: Change in Caudate Volume from Baseline

|  | Estimate | Std. Error | t value | p value |
| --- | --- | --- | --- | --- |
| (Intercept) | 1.8 | 0.49 | 3.6 | 0.0005 |
| antidepressant treatment | -0.23 | 0.026 | -8.6 | 1*.*2 *×* 10*^−^*^12^ |
| baseline caudate volume | -7 | 24 | -0.29 | 0.77 |

eTable 8c: Change in Putamen Volume

Estimate	Std. Error	t value	p value

| (Intercept) | 0.00013 | 0.00027 | 0.49 | 0.63 |
| --- | --- | --- | --- | --- |
| antidepressant treatment | -0.0001 | 1*.*4 *×* 10*^−^*^5^ | -7.4 | 2*.*4 *×* 10*^−^*^10^ |
| baseline putamen volume | -0.012 | 0.018 | -0.7 | 0.49 |

eTable 8d: Change in Whole Brain Volume

Estimate	Std. Error	t value	p value

| (Intercept) 2*.*3 *×* 10^2^ | 54 | 4.2 6*.*4 *×* 10*^−^*^5^ |
| --- | --- | --- |
| antidepressant treatment -11 | 2.7 | -3.9 0.00018 |
| baseline brain volume *−*1 *×* 10^2^ | 38 | -2.6 0.011 |

eTable 8e: Change in Grey Matter Volume

|  | Estimate | Std. Error t | value | p value |
| --- | --- | --- | --- | --- |
| (Intercept) | 52 | 1*.*3 *×* 10^2^ | 0.41 | 0.68 |
| antidepressant treatment | -13 | 4.3 | -3 | 0.0043 |
| baseline grey matter volume | *−*3*.*9 *×* 10^2^ | 1*.*3 *×* 10^2^ | -3 | 0.0042 |

eTable 9: Sensitivity Analysis - ENROLL-HD

|  | estimate | standard error | df | robustness (%) |
| --- | --- | --- | --- | --- |
| baseline composite score | 1.02 | 0.05 | 945.55 | 45.84 |
| visit year | -0.89 | 0.09 | 490.83 | 28.68 |
| antidepressant treatment: visit year | 0.36 | 0.12 | 479.02 | 4.98 |
| antidepressant treatment | -0.77 | 0.34 | 892.65 | 1.06 |
| CAG length | -0.19 | 0.09 | 938.51 | 0.33 |
| psychoactive drug | -0.82 | 0.40 | 929.77 | 0.30 |
| age | -0.02 | 0.02 | 945.86 | 0.00 |
| sex (male) | 0.02 | 0.02 | 936.32 | 0.00 |
| baseline pba depression score | 0.04 | 0.05 | 926.22 | 0.00 |
| baseline pba irritability score | 0.00 | 0.06 | 928.97 | 0.00 |
| baseline pba anxiety score | -0.05 | 0.05 | 924.03 | 0.00 |
| baseline pba suicidality score | 0.11 | 0.14 | 926.33 | 0.00 |
| number of antidepressant | -0.08 | 0.07 | 887.13 | 0.00 |
| previous mental health event | -0.26 | 0.32 | 935.62 | 0.00 |
| history of addiction | 0.18 | 0.32 | 933.84 | 0.00 |
| comorbidities | 0.33 | 0.35 | 921.21 | 0.00 |

eTable 10a: Antidepressant Effect on Composite Score Sequential Hotdeck Imputation

|  | Estimate | Std. Error | t value | p value |
| --- | --- | --- | --- | --- |
| (Intercept) | 10.37 | 5.16 | 2.012 | 0.044 |
| antidepressant treatment | -0.46 | 0.33 | -1.38 | 0.17 |
| visit year | -0.63 | 0.073 | -8.63 | *<* 2x10*^−^*^16^ |
| antidepressant treatment: visit year | 0.19 | 0.086 | 2.2 | 0.028 |

eTable 10b: Antidepressant Effect on UHDRS Motor Score Sequential Hotdeck Imputation

|  | Estimate | Std. Error | t value | p value |
| --- | --- | --- | --- | --- |
| (Intercept) | -8.99 | 29.74 | -0.3 | 0.76 |
| antidepressant treatment | 3.36 | 1.77 | 1.9 | 0.058 |
| visit year | 2.49 | 0.36 | 6.89 | 5.69x10*^−^*^12^ |
| antidepressant treatment: visit year | -0.96 | 0.46 | -2.087 | 0.037 |

eTable 10c: Antidepressant Effect on Neurofilament Light Chain Sequential Hotdeck Imputation

|  | Estimate | Std. Error | t value | p value |
| --- | --- | --- | --- | --- |
| (Intercept) | -46.85 | 38 | -1.2 | 0.22 |
| antidepressant treatment | -10.94 | 3.21 | -3.41 | 0.00086 |
| baseline neurofilament light chain | 0.044 | 0.11 | 0.41 | 0.69 |

eTable 10d: Antidepressant Effect on Change in Caudate Volume Sequential Hotdeck Imputation

|  | Estimate | Std. Error | t value | p value |
| --- | --- | --- | --- | --- |
| (Intercept) | 1.037 | 0.92 | 1.12 | 0.26 |
| antidepressant treatment | -0.18 | 0.071 | -2.56 | 0.011 |
| baseline caudate volume | -3.48 | 4.27 | -0.82 | 0.42 |

eTable 10e: Antidepressant Effect on Change in Whole Brain Volume Sequential Hotdeck Imputation

Estimate	Std. Error	t value	p value

| (Intercept) 2*.*8 *×* 10^2^ | | 51 | 5.6 1*.*4 *×* 10*^−^*^7^ | |
| --- | --- | --- | --- | --- |
| antidepressant treatment | -9.7 | 2.9 | -3.3 | 0.0011 |
| baseline whole brain volume | -89 | 31 | -2.9 | 0.0048 |
